# Cross-talk between the airway epithelium and activated immune cells defines severity in COVID-19

**DOI:** 10.1101/2020.04.29.20084327

**Authors:** Robert Lorenz Chua, Soeren Lukassen, Saskia Trump, Bianca P. Hennig, Daniel Wendisch, Fabian Pott, Olivia Debnath, Loreen Thürmann, Florian Kurth, Julia Kazmierski, Bernd Timmermann, Sven Twardziok, Stefan Schneider, Felix Machleidt, Holger Müller-Redetzky, Alexander Krannich, Sein Schmidt, Felix Balzer, Johannes Liebig, Jennifer Loske, Jürgen Eils, Naveed Ishaque, Christof von Kalle, Andreas Hocke, Martin Witzenrath, Christine Goffinet, Christian Drosten, Sven Laudi, Irina Lehmann, Christian Conrad, Leif-Erik Sander, Roland Eils

**Affiliations:** Center for Digital Health, Berlin Institute of Health (BIH) and Charité - Universitätsmedizin Berlin, corporate member of Freie Universität Berlin, Humboldt-Universität zu Berlin, Berlin, Germany; Molecular Epidemiology Unit, Charité - Universitätsmedizin Berlin, corporate member of Freie Universität Berlin, Humboldt-Universität zu Berlin, and Berlin Institute of Health (BIH), Berlin, Germany; Department of Infectious Diseases and Respiratory Medicine, Charité - Universitätsmedizin Berlin, corporate member of Freie Universität Berlin, Humboldt-Universität zu Berlin, and Berlin Institute of Health (BIH), Berlin, Germany; Institute of Viroloy, Charité - Universitätsmedizin Berlin, corporate member of Freie Universität Berlin, Humboldt-Universität zu Berlin, and Berlin Institute of Health (BIH), Berlin, Germany; Berlin Institute of Health (BIH), Berlin, Germany; Department of Tropical Medicine, Bernhard Nocht Institute for Tropical Medicine & I. Department of Medicine, University Medical Center Hamburg-Eppendorf, Hamburg, Germany; Sequencing Core Facility, Max Planck Institute for Molecular Genetics, Berlin, Germany; Clinical Study Center, Charité - Universitätsmedizin Berlin, corporate member of Freie Universität Berlin, Humboldt-Universität zu Berlin, and Berlin Institute of Health (BIH), Berlin, Germany; Department of Anesthesiology and Intensive Care Medicine, Charité - Universitätsmedizin Berlin, corporate member of Freie Universität Berlin, Humboldt-Universität zu Berlin, and Berlin Institute of Health (BIH), Berlin, Germany; Department of Anesthesiology and Intensive Care, University Hospital Leipzig, Leipzig, Germany; German Center for Lung Research (DZL), Germany; Health Data Science Unit, Heidelberg University Hospital and BioQuant, Heidelberg, Germany

## Abstract

The clinical course of COVID-19 is highly variable, however, underlying host factors and determinants of severe disease are still unknown. Based on single-cell transcriptomes of nasopharyngeal and bronchial samples from clinically well-characterized patients presenting with moderate and critical severities, we reveal the different types and states of airway epithelial cells that are vulnerable for SARS-CoV-2 infection. In COVID-19 patients, we observed a two- to threefold increase of cells expressing the SARS-CoV-2 entry receptor *ACE2* within the airway epithelial cell compartment. *ACE2* is upregulated in epithelial cells through Interferon signals by immune cells suggesting that the viral defense system may increase the number of potentially susceptible cells in the respiratory epithelium. Infected epithelial cells recruit and activate immune cells by chemokine signaling. Recruited T lymphocytes and inflammatory macrophages were hyperactivated and showed a strong interaction with epithelial cells. In critical patients, increased expression of *CCL2, CCL3, CCL5, CXCL9, CXCL10, IL8, IL1B* and *TNF* in macrophages was identified as a likely cause of a hyperinflammatory lung pathology. Moreover, we observed exacerbated epithelial cell death, likely leading to lung injury and respiratory failure in fatal cases. Our study provides novel insights into the pathophysiology of COVID-19 and suggests an immunomodulatory therapy along the CCL2, CCL3/CCR1 axis as promising option to prevent and treat critical course of COVID-19.

## INTRODUCTION

Since its outbreak in November 2019 in Wuhan, China, SARS-CoV-2 has spread rapidly worldwide with current disease hotspots mainly in Europe and in the USA with more than 2,883,603 WHO-confirmed SARS-CoV-2 infections worldwide and 198,842 deaths (as of 27 April 2020). While the median age of COVID-19 patients is 47 years, risk of developing severe illness increases sharply for patients aged 60 years and older [Guan *et al* (2020)]. No obvious sex prevalence for SARS-CoV-2 infection was reported, however, females might be slightly less severely affected [Guan *et al*. (2020)]. COVID-19 patients exhibit a broad spectrum of disease progression: 81% exhibit mild or moderate symptoms with very few asymptomatic cases, 14% show severe symptoms and 5% critical disease progression. [Wu & McGoogan (2020)].

Coronaviruses, including SARS-CoV-2, infect and replicate in both the upper and lower respiratory tract. The upper airways are specialized in eliminating inhaled pathogens to prevent viral invasion in the lower respiratory tract and subsequent pulmonary injury [Branchett & Lloyd (2019)]. Upon virus infection, epithelial cells interact with immune cells to control viral spread. These interactions are largely mediated by cytokine signaling and cell-cell contacts. After successful viral clearance, restoration of homeostasis by controlled elimination of activated immune cells is necessary in order to avoid hyperactivation of the immune system and exacerbated tissue damage [Branchett & Lloyd (2019), Tisoncik *et al* (2012)]. It has been suggested that severe lung injury observed in some critical COVID-19 patients is rather a consequence of the hyperactivated immune system than of the disability of viral clearance [Mehta *et al* (2020), Wen *et al* (2020)]. However, mechanistic insights supporting this hypothesis are still missing. Here, we present a comprehensive study of mucosal immune responses in COVID-19 patients that aims at understanding the cellular and molecular basis of different severities of disease. This study is based on single cell RNA sequencing data performed on samples from the respiratory system of moderate, critical and in the clinical course deceased COVID-19 patients, including longitudinal and multiple sampling along the respiratory tract.

## RESULTS

### Cellular and molecular characterization of COVID-19 severity

To dissect the cellular and transcriptional heterogeneity in the respiratory tract of COVID-19 patients with a moderate and critical course of disease, respectively, we performed 3’ single cell RNA sequencing (scRNAseq) on nasopharyngeal or pooled nasopharyngeal/pharyngeal swabs (NS), bronchiolar protected specimen brushes (PSB), and bronchoalveolar lavages (BAL) (Figure 1a). We analyzed a single-hospital cohort from Charité - Universitätsmedizin Berlin consisting of 14 patients (ten males, four females; 21 to 75 years of age; median 52.5 years; Figure 1b, Extended Data Table 1). According to the WHO guidelines [Aylward *et al* (2020)], five patients were classified as moderate and nine patients as critical with two of them deceased in the clinical course (Figure 1b, patients #15, #32). Four COVID-19 patients were sampled longitudinally to investigate cellular and transcriptional dynamics during the course of disease (patients #1, #5, #7, #8). For two patients, multiple samples were collected (NS, PSB, BAL) to compare cellular and transcriptional heterogeneity in the upper and lower airways (patients #32, #62). In addition, we investigated one Influenza B patient (patient #4) who was hospitalized with severe symptoms. He was tested negative for SARS-CoV-2 but positive for influenza-B. Two SARS-CoV-2 negative controls from supposedly healthy volunteers (both male, 24 and 51 years; for further detail see Methods) were included as well. Based on transcriptional profiles of 168,596 cells obtained from a total of 29 samples taken from 17 individuals, we determined the cellular landscapes of COVID-19, Influenza B and control patients (Figs. 2a and b; Extended Data Fig. 6). In COVID-19 nasopharynx samples we identified 21 different cell states spread between epithelial cells and immune cells (Figs. 2a and b). We detected all previously described major epithelial cells of the conducting airways, including basal, secretory, and ciliated cells, as well as the recently discovered FOXN4+ cells and ionocytes (Methods, Figs. 2a and b, Extended Data Fig. 1a, Suppl. Table 1) [Montoro *et al* (2018), Plasschaert *et al* (2018), Vieira Braga *et al* (2019)]. Interestingly, a subpopulation of basal cells showed a distinctively strong Interferon gamma (IFNG) response signature (IFNG mediated basal cells annotated as basal INF), which is atypical for resting basal cells and points towards their stimulation by the host immune system (Fig 2a, Extended Data Fig. 1a) [Schneider *et al* (2014), Newton *et al* (2016)]. Within the immune cell population, we identified twelve different cell types, belonging to macrophages, dendritic cells, mast cells, neutrophils, B cells and T cells (Fig 2a and b, Extended Data Fig. 1b, Suppl. Table 1; for further details see Methods). Surprisingly, we were not able to identify NK cells, usually characterized by the presence of *NCAM1* and absence of *CD3E*, in the nasopharyngeal samples. Such a population was only present in the samples of the lower respiratory tract (see below).

**Figure 1:**
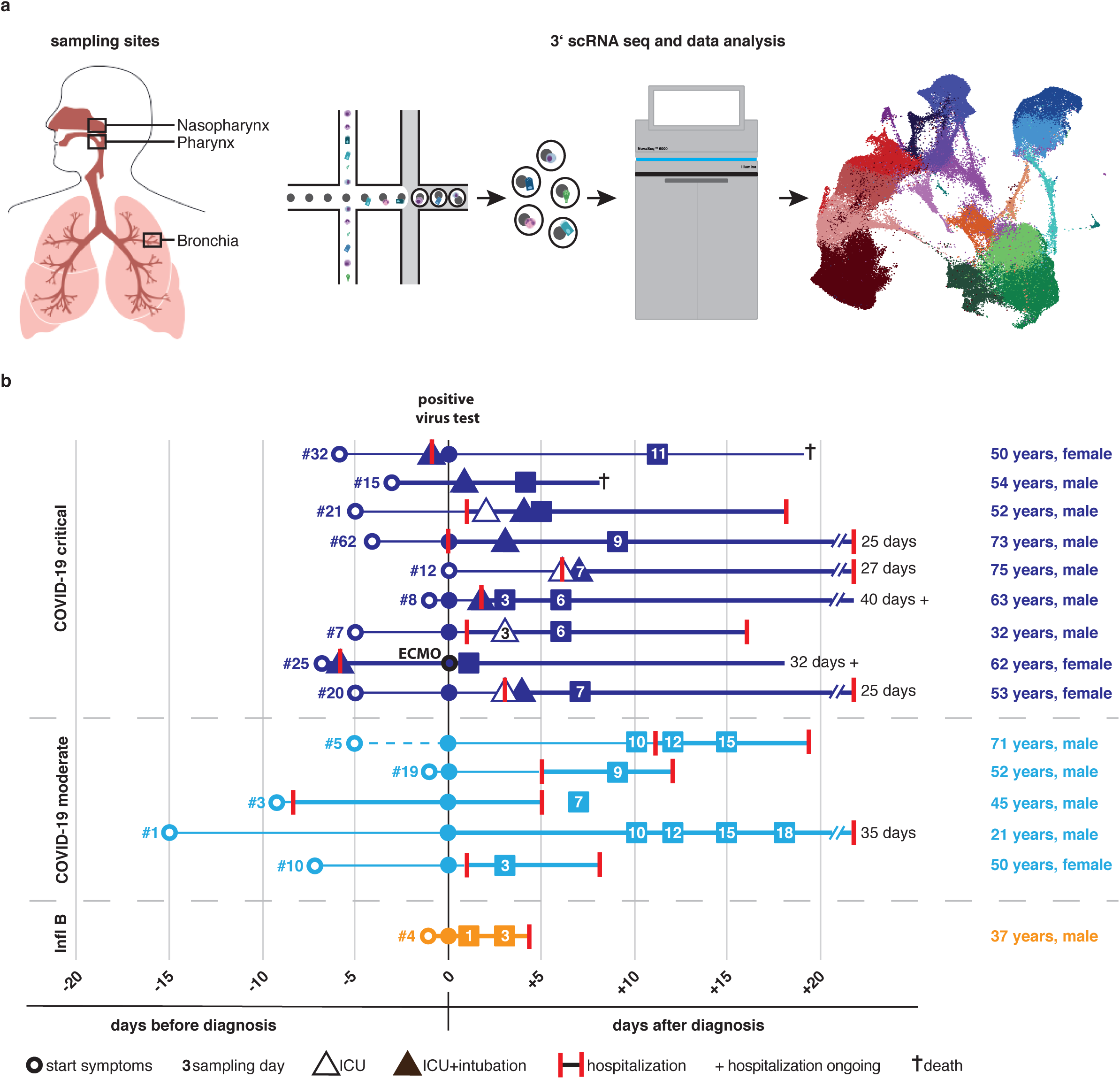
Illustration of the experimental setup and the patient cohort used in this study. **A**. Schematic representation of the experimental workflow: Depicted are the sampling sites (left) and the 3’ scRNAseq library preparation using 10x Genomics (middle) followed by data analysis revealing cell type identification (right). **B**. Overview of patient cohort depicting age, sex, classification of COVID-19 severity as well as onset of diseases, hospitalization duration and sampling time points, with all patients being temporally aligned to the day of diagnosis.

**Figure 2.**
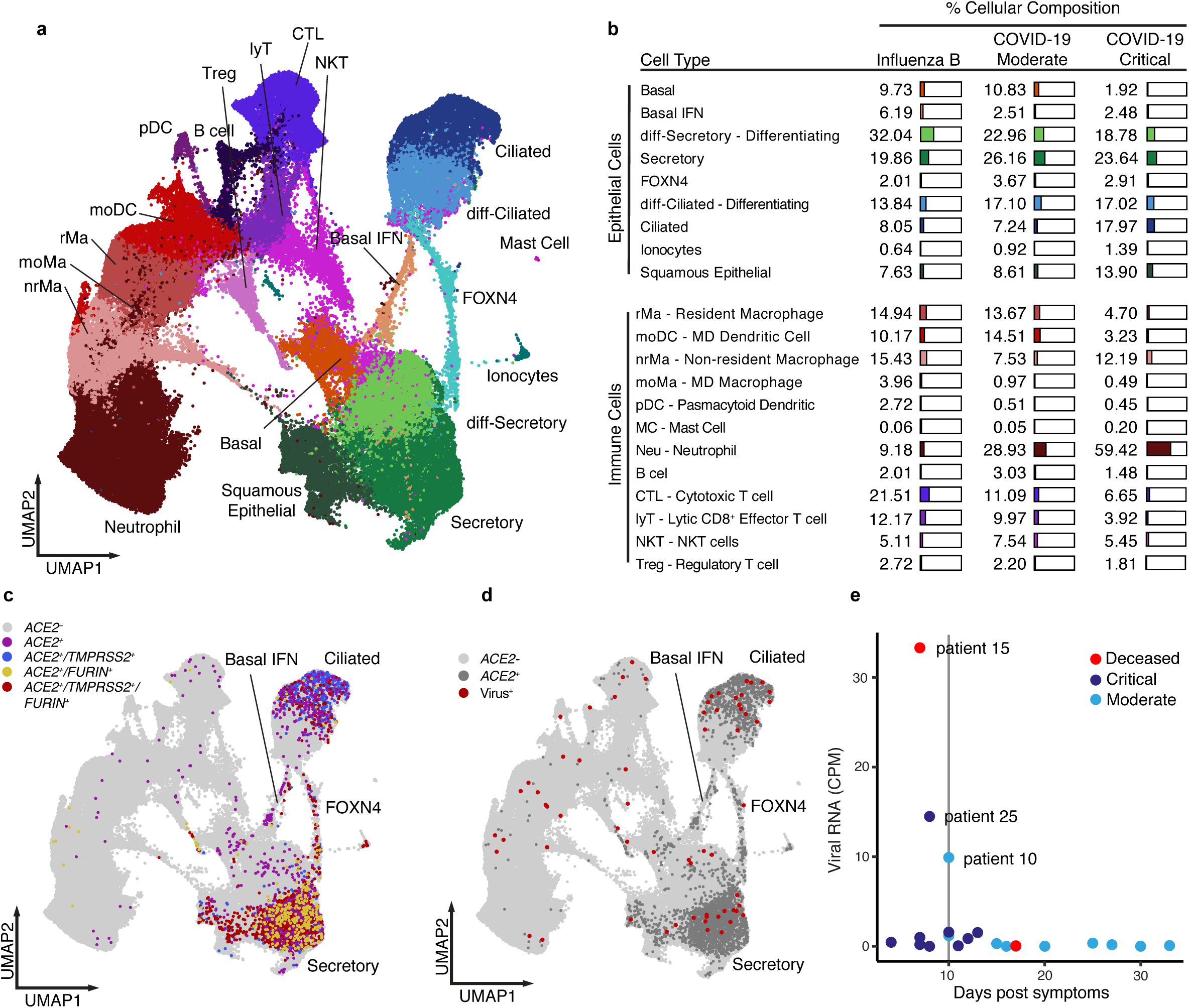
Identification, distribution and classification of cells from the upper respiratory tract. **A**. UMAP displaying all identified cell types and states. **B**. Table displaying the total contribution of each cell type aggregated for Influenza B, COVID-19 moderate, and COVID-19 critical cases in percentage of total epithelial and immune cell types, respectively. **C**. Distribution of *ACE2^+^, ACE2^+^/TMPRSS2*^+^, *ACE2^+^/FURIN^+^, ACE2^+^/TMPRSS2^+^/FURIN^+^* cells across all cell types within the UMAP. **D**. UMAP depicting *ACE2^+^* cells (dark grey) and virus RNA+ cells (red). **E**. Viral RNA reads within aggregated pseudo-bulks for each patient sample in counts per million (CPM) against days post symptoms. Note that only very low numbers of viral RNA reads were detected 10 days after first symptoms and later.

Comparing the cellular landscape of moderate versus critical COVID-19 patients (Fig. 2b, Suppl. Table 1), we observed a striking depletion of basal cells (p-value <2.06E-6) and strong enrichment for neutrophils (p-value < 2.18E-71) in critical COVID-19 patients (Fig. 2b, Suppl. Table 1). These changes should be carefully interpreted since the nature of an active viral infection is very dynamic and populations of cells change dramatically over the course of a viral infection [Ibricevic *et al* (2006), Lam *et al* (2008), Steuerman *et al* (2018)]. Moreover, clinical and genetic differences between patients, date of sampling, and the days since the onset of symptoms, further impact the variability of the cellular landscape of patients.

### Variable *ACE2* expression in the upper airway epithelium

SARS-CoV-2 is thought to initially infect the mucosa of the upper respiratory tract, facilitated by viral binding to the host receptor ACE2 and S protein priming by specific proteases, such as TMPRSS2 or FURIN [Hoffmann *et al* (2020), Walls *et al* (2020), Yan *et al* (2020), Zhou *et al* (2020), Coutard *et al* (2020)]. Co-expression levels of *ACE2* with either one or both S-priming proteases exceeded those detected in the non-infected control samples by two- to three-fold, specifically in secretory and ciliated cells (Fig. 2c, Extended Data Fig. 2c and d, Suppl. Table 2). Most ciliated cells were *ACE2^+^/TMPRSS2^+^*, while secretory and FOXN4^+^ cells were predominantly *ACE2^+^*/*TMPRSS2*^+^/*FURIN^+^* (Fig. 2c, Extended Data Fig. 2d, Suppl. Table 2). Secretory and ciliated cells also contained the highest fraction of SARS-CoV-2-infected cells (Fig. 2d). In agreement with a recent study by Wolfel *et al* (2020), the number of detected viral transcripts as count per million (CPM) reads was rather low in all COVID-19 patients and only detectable at considerable levels until ten days after onset of symptoms (Fig. 2e), which is in agreement with a recent study showing that infectious virus shedding disappeared 9–10 days post symptom onset [Wolfel *et al*. (2020)]. Note that the enrichment of *ACE2^+^* cells was significantly overrepresented within the epithelial cells compared to the immune cells (29 to 45-fold enrichment, p < 0.005). Viral transcripts captured in a very low number of immune cells are likely caused by viral association, e.g. phagocytosis or surface binding of viral particles, rather than productive infection (Fig. 2d). This would be in line with a study reporting virtual absence of viral reads in peripheral blood mononuclear cell (PBMC) samples from COVID-19 patients [Xiong *et al* (2020)].

### Dynamics of epithelial differentiation upon SARS-CoV-2 infection

As the stem cells of the airway epithelium, basal cells (*TP63^+^/KRT5*) preferentially differentiate into secretory cells that further differentiate into ciliated cells [Pardo-Saganta *et al* (2015)]. This differentiation trajectory changes during an insult or injury of the airway epithelium. In the distal airways of the lung, basal cells were found to be the main drivers of the wound healing process, as they undergo proliferative expansion and subsequently differentiate into ciliated and secretory cell types [Pardo-Saganta *et al*. (2015), Vaughan *et al* (2015), Zuo *et al*(2015), Montoro *et al*. (2018)].

Here, we inferred differentiation paths of basal cells from single-cell sequencing data of COVID-19 patients by pseudotime mapping using reversed graph embedding (Extended Data Fig. 3a) [Qiu *et al* (2017)]. The prototypical differentiation path from basal cells, through secretory to terminally differentiated ciliated cells is mediated by FOXN4^+^ cells (Extended Data Fig. 3a and b). Genes driving this differentiation path include the master transcription factor for ciliated cells, *FOXJ1*, and a component for mature cilia, *TCTEX1D2* [Yu *et al* (2008), Cruz *et al* (2010)]. Besides this classical path, we also predicted an alternative differentiation path leading from basal cells directly into ciliated cells. This direct differentiation path is dependent on IFNG mediated basal cells mostly driven by interferon response genes (ISGs) like *ISG15, IFIT1, IFIT3*, and *IFITM3* (Extended Data Fig. 3c) [Rusinova *et al* (2013)]. The hitherto unknown dependency of this differentiation path on interferon response to viral mediated injury resembles the recently described direct differentiation path from basal to ciliated cells upon chemical injury [Montoro *et al*. (2018)].

### Immune cell - epithelial interactions increase infectability of epithelial cells

A puzzling hallmark of SARS-CoV-2 infection is the surprisingly small number of *ACE2^+^* target cells within the human respiratory tract of healthy controls [Lukassen *et al* (2020), Sungnak *et al* (2020)]. Upon SARS-CoV-2 infection, *ACE2* expression levels increase by two- to three-fold in COVID-19 patients (Fig. 2c, Extended Data Fig. 2c, Suppl. Table 2). Interestingly, we observed a strong and statistically significant correlation between the percentage of both *ACE2^+^* ciliated and secretory cells and the intercellular-signaling strength of those cells with cytotoxic T lymphocytes (CTL) (Fig. 3b, see methods section for further details). *ACE2* expression has been identified as interferon-responsive [Ziegler *et al* (2020)], and *STAT1*, a main transcription factor of interferon response [Krause *et al* (2006)], was among the top predictors for *ACE2* expression (Extended Data Fig. 4a). The preferential expression of *IFNG* by CTLs and genes encoding its receptor *IFNGR1/2* by secretory and ciliated cells supports the notion that *ACE2* is upregulated in epithelial cells at least partially through IFNG signaling by immune cells (Extended Data Fig. 4b). Thus, this upregulation of *ACE2* as an interferon-stimulated gene (ISG) may counteract viral infection in general [Rodrigues Prestes *et al* (2017)]. In COVID-19 patients, however, an increase in the number of *ACE2^+^* cells potentially renders these patients even more vulnerable to SARS-CoV-2 infection. It is thus tempting to speculate that the defense of CTLs against SARS-CoV-2 infection may also play a key role in increasing the vulnerability of the airway epithelium to infection. Together, with the above described *IFNG*-driven direct differentiation from basal to largely *ACE2^+^* ciliated cells in COVID-19 patients, this host response pathway potentially increases the number of *ACE2^+^* cells and thus susceptibility of the airway epithelium to the SARS-cov-2 virus.

**Figure 4.**
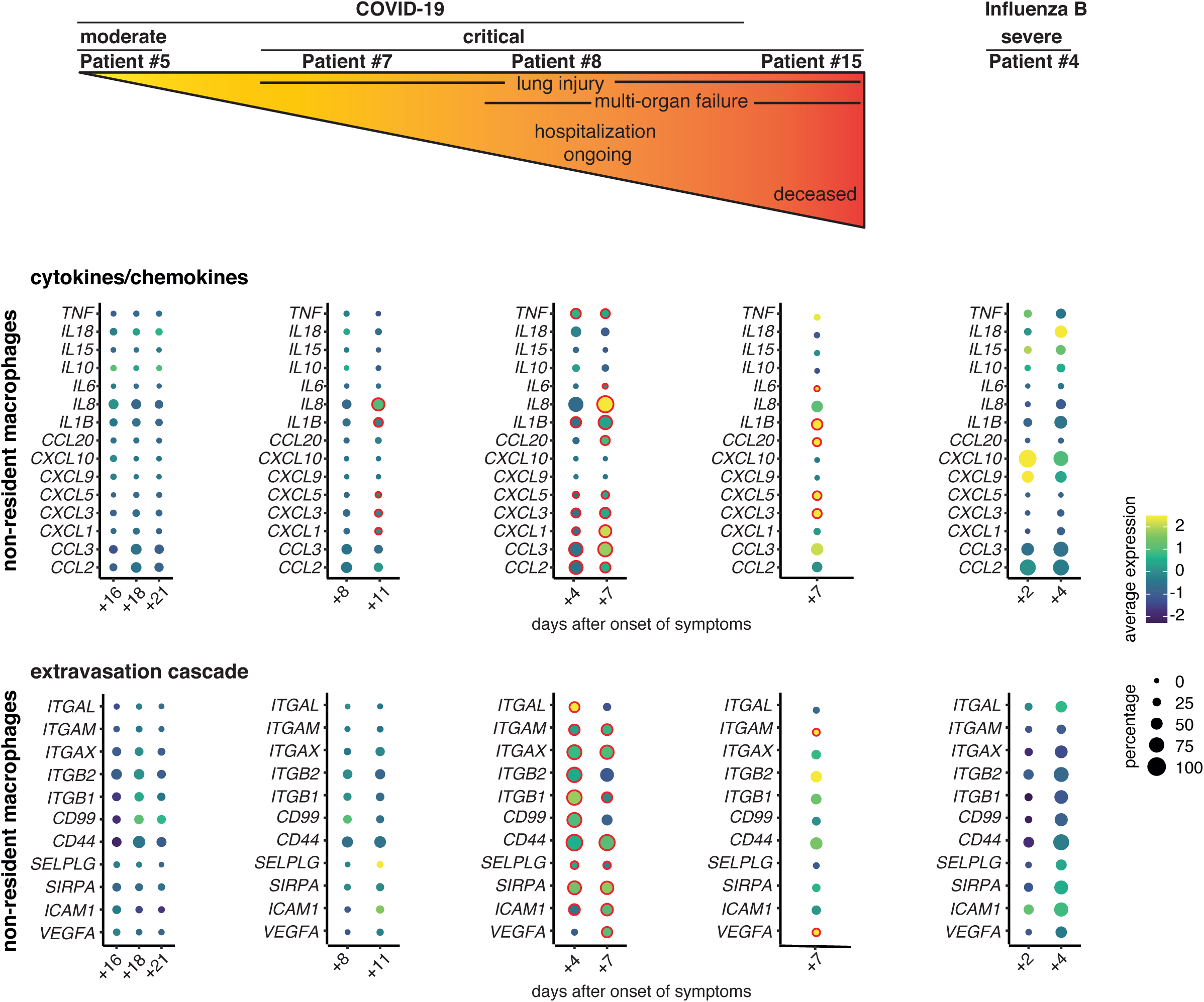
Differences between moderate and critical COVID-19 severity is attributed to non-resident macrophage extravasation and over-expression of cytokines/chemokines. Dot plots depicting the average expression of specific cytokines/chemokines and extravasation cascade genes of moderate and critical COVID-19 patients, and a severe Influenza B patient. COVID-19 patients are ordered by severity as indicated by different clinical features. Different times of sampling per patient are reflected by “days after onset of symptoms.” Expression levels are color-coded, the percentage of cells expressing the respective gene is size-coded. Significant increases for each time point and patient versus an average of the three time points of the moderate patient #5 (left panel) are marked by a red circle. Note that no statistical analysis for differences across the two diseases was performed. The proportion of cells expressing markers of the extravasation cascade were slightly increased in the Influenza B patient at the second time point; however, the expression level of these genes did not change.

### Immune-mediated epithelial damage in severe COVID-19

Infected epithelial cells were found to secrete chemokines that recruit and activate different immune cell populations at the site of viral infection (Extended Data Fig. 5). In moderate COVID-19 patients, secretory cells strongly expressed chemokine ligand encoding genes *CXCL1, CXCL3, CXCL5*, and *CXCL16*, which promote the recruitment of neutrophils and CTLs, respectively (Extended Data Fig. 5, upper panel). Ciliated cells expressed *CCL15* which may contribute to an inflow of monocytes/macrophages and neutrophils via CCR1. Epithelial-derived chemokine encoding genes, such as *CXCL6, CXCL16, CXCL17, and CCL28*, were not further assessed, as the immune cells of the COVID-19 patients did not express the respective receptors (Extended Data Fig. 5). The chemokine and chemokine receptor expression of the different cell populations increased remarkably in the critical compared to the moderate cases, pointing towards an augmented recruitment of immune cells to inflamed sites (Extended Data Fig. 5, lower panel).

In order to map the resulting complex interaction of epithelial and immune cells, we inferred all possible intercellular communications by the expression of multi-subunit ligand-receptor complexes in both cell populations with CellPhoneDB (Fig. 3a) [Efremova *et al* (2020)]. In moderate cases, the strong interaction between ciliated cells, secretory cells, CTL, non-resident macrophages (nrMa), monocyte-derived macrophages (moMa), and neutrophils may reflect the necessary immune response for the elimination of SARS-CoV-2 infected epithelial cells (Fig. 3a left). Interactions between the different cell types increased in the critical COVID-19 patients compared to the moderate cases (Fig. 3a right). Increased numbers of epithelium-immune cell interactions in critical COVID-19 cases are consistent with a higher activation status of nrMa, moMa, and CTL (Fig. 3c). In particular, nrMa showed a highly inflammatory profile characterized by the expression of the chemokine encoding genes *CCL2 (MCP1), CCL3 (MIP1α), CCL20, CXCL1, CXCL2, CXCL3, CXCL5*, and the pro-inflammatory cytokines *IL1B, IL8, IL18*, and *TNF*. Except for *CCL3* and *IL1B*, each of these inflammatory mediators was significantly higher expressed in critical COVID-19 cases (Fig. 3c). moMa were mainly characterized by *CXCL9* and *CXCL10* expression in the moderate COVID-19 patients, and additionally high *CCL2* and *CCL3* expression levels in the critical cases (Fig. 3c). Notably, increased *CCL2* and *CCL3* expression corresponded to an induction of *CCR1*, the receptor of *CCL2* and *CCL3*, in neutrophils and different macrophage populations (Fig. 3g). It has been repeatedly suggested that an excessive production of pro-inflammatory cytokines promotes the development of fatal pneumonia and acute lung injury in COVID-19 [Mehta *et al*. (2020), preprint: Wen *et al*. (2020)]. CCL2 and CCL3 may contribute to excessive inflammation by promoting monocyte recruitment, and increased levels of those chemokines were found in plasma samples of COVID-19 patients with a fatal outcome [Huang *et al* (2020)].

In the moderate cases, CTL displayed the characteristic transcriptional profile of anti-viral *CD8*^+^T lymphocytes with high expression of *IFNG*, *TNF*, *CCL5*, *PFR1* (encoding perforin) and *GZMB*, *GZMA* (encoding granzymes) together with genes encoding for cytotoxic receptors (*KLRB1, KLRC1, KLRC2, KLRD1*) (Fig. 3c). In the critical COVID-19 patients, CTL expressed only low levels of *IFNG* and *TNF* but strongly increased their cytotoxic potential (Fig. 3c). Cytotoxic T cells play a key role in virus elimination, but unrestrained CTL activity can cause excessive epithelial damage and lung injury [Xu *et al* (2004), Connors *et al* (2016)]. The result of such a damage might be reflected by the high levels of *CASP3* expression in ciliated, secretory and FOXN4^+^ cells (Fig. 3d).

**Figure 3.**
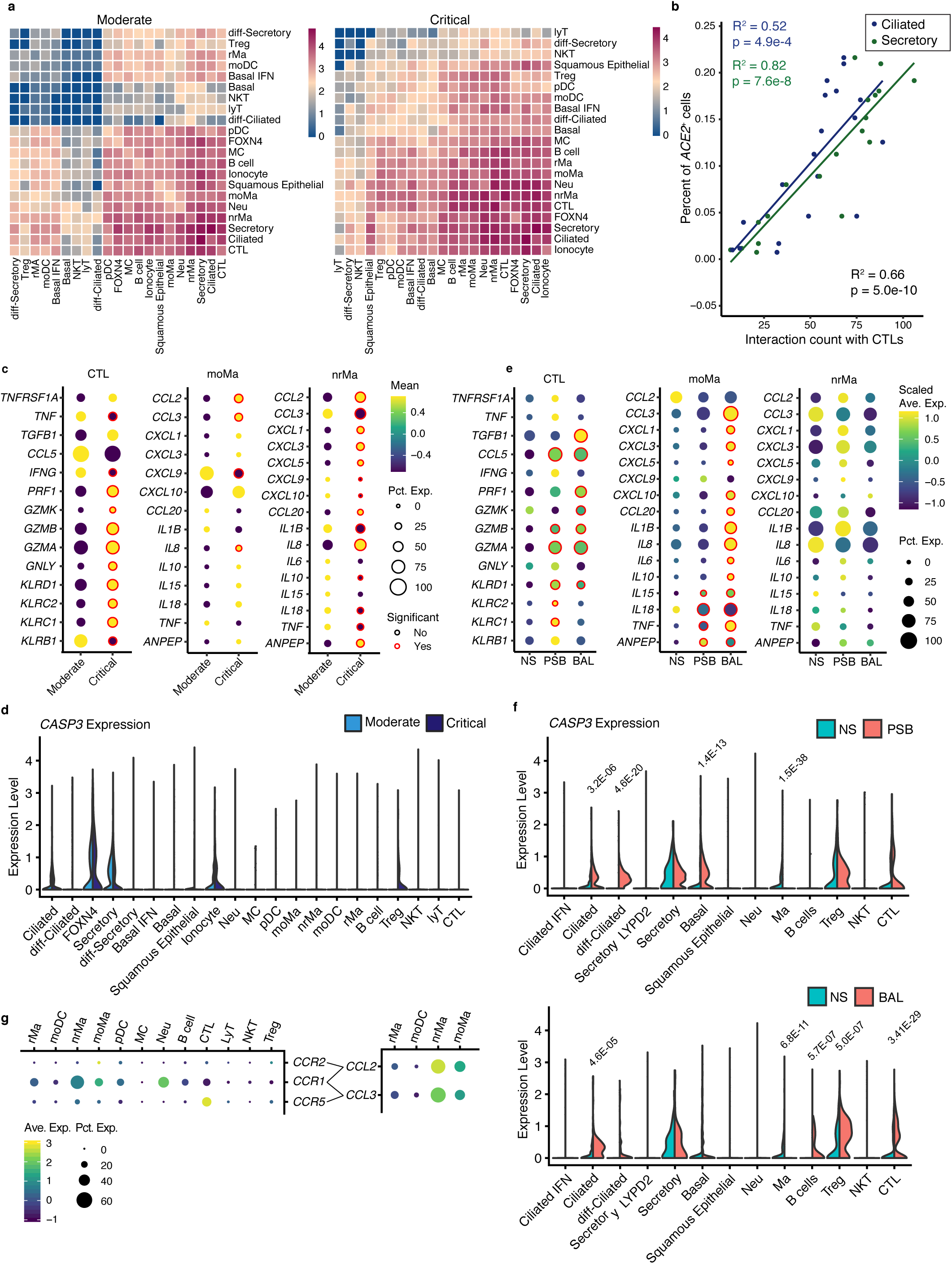
Immune – epithelial cell interaction in COVID-19. **A**. Heat map depicting cell-cell communications between all identified epithelial and immune cell types derived from log-scaled ligand-receptor interaction counts in moderate (left, n=5 patients, 10 samples) and critical excluding fatal (right, n=6 patients, 10 samples) COVID-19 patients. Interaction strengths as predicted by CellPhoneDB are color-coded. **B**. Scatter plot displaying the expression level of *ACE2* in secretory and ciliated cells vs the interaction strength of the respective cell type with CTLs, measured by number of expressed ligand-receptor pairs, in COVID-19 patients (n=12 patients, 19 samples). Cell type is color-coded, R^2^ and p-values indicated for CTL interaction with secretory cells (green), ciliated cells (blue), and combined ciliated and secretory cells (black), respectively. **C**. Dot plots depicting the expression of pro-inflammatory and cytotoxic mediators in CTL, moMa and nrMa. Expression levels are color-coded, the percentage of cells expressing the respective gene is size-coded, and significantly increased expression in critical (excluding fatal cases) versus moderate is marked by a red circle. D. Comparison of *CASP3* expression in the different epithelial and immune cell types of critical and moderate COVID-19 cases (n=5 moderate, 6 critical). **E**. Dot plots showing expression changes of inflammatory and cytotoxic mediators from nasal swabs (NS), protected specimen brush samples (PSB) and bronchioalveolar lavages (BAL) collected from the same individual. Data shown here is taken from two critical patients #32 and #62. Expression levels, percentage of cells, and significance levels visualized like in C above. Note no significance was reached for nrMa. **F**. Comparison of *CASP3* expression in the different immune and epithelial cell subtypes in PSB vs. NS (top) and BAL vs. NS (bottom) for patients #32 and #62. P-values for significant decrease in nasal vs. other sampling sites are depicted above cell type (missing value indicate no significance) **G**. Dot plot depicting the average expression of *CCL3/CCR1, CCR5* and *CCL2/CCR2* in the different immune cells. In A-D and G, an average of n = 5 moderate and n = 6 critical patients with 10 samples per moderate/critical group is shown.

To assess whether the inflammatory macrophages and pathogenic CTL observed in the nasopharyngeal mucosa were also present in the lower airways, we collected multiple samples of two patients (#32 and #62) from the upper (NS) and lower airways (PSB and BAL). When integrating the scRNAseq data from these triplets of samples for the two patients, we observed that all epithelial and immune cell populations were present in all three sampled sites of the airways (Extended Data Fig. 6a and b). Further, distribution of *ACE2^+^* cells was spread across all three sites, whereas cells containing viral reads primarily appeared in neutrophils in BAL samples, potentially reflecting the phagocytosis of apoptotic virus-infected epithelial cells (Extended Data Fig. 6c). This is in agreement with our observation that the viral load as measured by total counts of viral reads and viral copy numbers were higher in BAL compared to the nasopharyngeal mucosa for both patients (Extended Data Fig. 6d-e).

Macrophages in the lower airways (PSB and BAL) had a higher inflammatory potential than those within the upper airway (NS) as both the expression level for genes encoding for all the inflammatory chemokines/cytokines and the number of macrophages expressing them were significantly elevated (Fig. 3e). These data are in agreement with a preliminary study that performed scRNAseq on BAL cells showing an inflammatory macrophage phenotype in COVID-19 patients expressing *CCL2, CCL3, IL8, CXCL9*, and *CXCL10* [preprint: Liao *et al* (2020)]. The presence of highly inflammatory macrophages and cytotoxic T cells (Fig. 3e) corresponds with a high *CASP3* expression in the PSB as well BAL samples. Interestingly, the potential for apoptosis in basal and ciliated cells is significantly higher in the lower (PSB or BAL) compared to the upper airways (NS) reflecting the presence of highly inflammatory macrophages and CTL (Fig 3f).

### Patient-specific immune inflammation profiles

To systematically compare the pronounced differences in the cytokine/chemokine expression profiles between moderate and critical COVID-19 cases at the individual patient level, we compared expression pattern within non-resident macrophages in longitudinal samples of patients with different degrees of disease severity (Fig. 4). Patient #5 (male, 71 years old) is a typical moderate case of COVID-19. Patient #7 - a middle-aged male without any risk factors - is a critical case of COVID-19 with transient need for high flow oxygen ventilation. He was admitted to the ICU with an isolated respiratory failure due to bipulmonary infiltrates. He was discharged from ICU after two days, and left the hospital in good health. Patient #8 is a 63-year-old male, obese with a history of smoking, and he represented a critical case of COVID-19. He required mechanical ventilation and developed severe acute respiratory distress syndrome (ARDS) and multi-organ failure. After two weeks in the ICU his condition gradually improved and he could be extubated. Patient #15 is a critical case of COVID 19 with severe ARDS and fatal outcome.

The expression patterns of genes encoding for cytokine/chemokine between patient #5 and #7 were similar. Only a slight increase for *IL8* expressing non-resident macrophages over time was observed for patient #7 (Fig. 4). Both mechanically ventilated critical cases (#8 and #15) showed increased expression of *CCL20, CXCL5, CXCL3, CXCL1, CCL3*, and *CCL2* along with an enhanced expression of *TNF* and *IL1B* compared to the patients #5 and #7 (Fig. 4). In patient #8, these changes increased over time converging on a similar pattern as observed for patient #15 who deceased shortly after the depicted observation time point. This would be in line with the clinical course of patient #8, as he remained in critical condition ill for a prolonged period of time.

Extravasation of immune cells, the recruitment of blood monocytes to the infected or injured tissue and their migration across the endothelium are crucial events in early immune defense of infected epithelial cells, but they are also critically involved in capillary leakage and lung damage. Although the primary site of infection by SARS-CoV-2 seem to be epithelial cells, reports are emerging that also the endothelium is infected by SARS-CoV-2 *in vitro* [Monteil *et al* (2020)] and *in vivo* [Varga *et al* (2020)]. Endothelial inflammation has been observed in COVID-19 patients and was associated to endothelial dysfunction and an impaired microcirculation, both hallmarks of ARDS and other clinical sequelae of COVID-19 (e.g. acute renal failure).

A variety of molecules is involved in monocyte trafficking across the vessel wall (e.g. integrins) [Gerhardt & Ley (2015)]. Comparing the expression of markers along the extravasation cascade with respect to COVID-19 disease severity, we observed a strong increase of these genes in the non-resident macrophages. *ITGAL (CD11a), ITGAM(CD11b), ITGAX* (CD11c) *ITGB1 (CD29)* and *ITGB1 (CD18)* encode for integrins forming the complexes LFA-1, Mac-1 and VLA-4 that are all described as essential for monocyte migration across the endothelium [Meerschaert & Furie (1995), Gerhardt & Ley (2015)].

Patient #8 showed a significant increase of *ITGAM, ITGAX, ITGB2* and *ITGB1* expressing cells at both time-points investigated compared to the moderate patient #5 (Fig. 4). This is in line with the increased proportion of *CD44*, a crucial mediator of host defense, expressing cells in this patient [Rivadeneira *et al* (1995), Vachon *et al* (2007), Xu *et al* (2008), He *et al* (2016)]. We found *ICAM1* and *VEGFA*, attractant factors commonly expressed by endothelial (and epithelial) cells to recruit leucocytes to sites of infection, also expressed by monocytes themselves, suggesting that activated monocytes induced their own recruitment. The increased expression of these extravasation markers indicates an enhanced anti-viral immune response, which might damage the epithelial and endothelial barrier alike.

Patient #15, with the most critical clinical course, showed a similar pattern of extravasation marker expression; however, these changes were only significantly increased for *ITGAM* and *VEGF*. In general, a smaller proportion of non-resident macrophages expressed the extravasation markers in this patient, which might be the reason why other marker gene expression differences were not significant.

Patient #4 was hospitalized as a symptomatic contact person for suspected COVID-19, but was tested negative for SARS-CoV-2 and positive for influenza B. The proportion of non-resident macrophages expressing chemokines/cytokines at a high level decreased over time, and the cellular patterns and expression of extravasation genes are visually distinct from the ones in critical COVID-19, corresponding to the discharge of this patient shortly thereafter.

### Potential therapeutic implications

The data shown in our manuscript support the well-known role of ACE2, which places the receptor and its co-factors as primary drug target. However, from a clinical perspective the main determinant for different clinical courses of COVID-19 infections is a hyperactivated, pathogenic immune response. It is hypothesised that especially an exaggerated immune response causes the severe and critical COVID-19 courses with a potentially fatal outcome. Therefore, immune modulators should be considered as a primary therapeutic principle for treatment of COVID-19. The current focus in development of COVID-19 therapies is based on inhibition of immunological key players like IL6, TNF or interferon [Coomes & Haghbayan (2020)] (NCT04359901, NCT04322773, NCT04311697). However, our data suggest that targeting chemokine receptors could also offer a promising therapeutic option. We find a significant induction of *CCL2* and *CCL3* expression in macrophages corresponding with an increased expression of *CCR1*, the receptor for both chemokines, in critical COVID-19 patients. Since binding of *CCL2* or *CCL3* to *CCR1, CCR2 and CCR5* can induce monocyte recruitment into the lung parenchyma with consecutive recruitment and activation of further immune cells and epithelial damage, *CCR1, CCR2, and CCR5* might represent promising anti-inflammatory targets in COVID-19. In HIV and other viral infections targeting the CCR2/CCL2 axis was already introduced as a potential therapeutic strategy [Covino *et al* (2016)]. However, since we observed *CCR2* expression neither in the upper nor in the lower respiratory tract of COVID-19 patients (Extended Data Fig. 7), CCR1 inhibition, possibly combined with CCR5 blockage, might be a potentially more promising strategy in SARS-CoV-2 infection.

## METHODS

### Patient Recruitment and Ethics Approval

All patients were enrolled in the prospective observational Cohort Study (Pa-COVID-19) at Charité - Universitâtsmedizin Berlin. We obtained a signed informed consent for inclusion in this study. The study was approved by the institutional ethics committee of the Charité - Universitâtsmedizin Berlin (EA2/066/20) and conducted in accordance with the Declaration of Helsinki.

### Patient cohort

All COVID-19 patients described common cold symptoms like dry cough and fever, followed by malaise, cephalalgia and myalgia at the onset of the disease. In addition, dyspnea was a common start symptom among the critical cases. The median duration from first symptoms until hospital admission was 8 days for the moderate cases (range: 1–17 days) and 5 days for the critical cases (range: 1–6 days). Moderate COVID-19 patients were hospitalized for a median duration of 8 days, while critical cases were hospitalized for a median duration of 25 days with some patients being hospitalized for more than 40 days. Most of the critical COVID-19 patients required mechanical ventilation within the first 8 days after the onset of symptoms with a median duration of the intensive care unit (ICU) stay of 25 days. The only exception was patient #7, who only stayed for two days in ICU with nasal high-flow oxygen ventilation. Patient #25 was the only patient in this study cohort that required extracorporeal membrane oxygenation (ECMO) support. The sepsis-related/sequential organ failure assessment (SOFA) scores as well as the ARDS scores of the ICU patients were used as indicator for multi-organ dysfunction or lung injury, respectively (see Extended Data Table 1). Sex, age, and information regarding the COVID-19 onset of symptoms, diagnosis and hospitalization for each individual is provided in Figure 1b and Extended Data Table 1. Two critical COVID-19 patients (#15 and #32) deceased.

In addition to the COVID-19 patients, our cohort included one Influenza B patient (#4, male, 37 years) with respiratory symptoms (cough, dyspnea) and a hospitalization duration of four days. Further, two supposedly healthy volunteers (both male, 24 and 51 years) from SARS-CoV-2 negative controls were included. Based on our scRNAseq data analysis, it appears that one of the control donors (ctrl #2) was recovering from a viral infection. Note that the here presented cohort does not represent the patient distribution admitted to Charité with regards to sex, age, or COVID-19 severity as the patients were randomly chosen based on their presence in the hospital and willingness to donate samples for this study.

### Isolation and preparation of single cells from human airway specimens

We used human airway specimens freshly procured using nasopharyngeal swabs, bronchial brushes, or bronchioalveolar (BAL) lavages for single-cell RNA. The nasopharyngeal swabs, and bronchial brushes taken from donors were directly transferred into 500 μL cold DMEM/F12 medium (Gibco, 11039). Processing of all samples started within an hour after collection and under biosafety S3 conditions (non-infectious donor nasopharyngeal samples were processed under biosafety S1/S2 conditions).

Nasopharyngeal and bronchial samples were pre-treated with 500 μL of 13mM DTT (AppliChem, A2948), followed by dislodging the cells from the swabs/brushes by carefully pipetting the solution onto the swab/brush. The swab/brush was dipped 20 times in the solution to ensure the release of all cells and then discarded. BAL samples were immediately treated with an equal volume of 13mM DTT. Afterwards, all following steps were identical for nasopharyngeal, bronchial, and BAL samples: All samples were incubated on a thermomixer at 37°C, 500 rpm for 10 minutes and then spun down at 350xG at 4°C for 5 minutes^1^. The supernatant was carefully removed, followed by the inspection of the cell pellet for any trace of blood. If so, cells were resuspended in 500 μL 1x PBS (Sigma-Aldrich, D8537) and 1 mL of RBC Lysis Buffer (Roche, 11814389001), incubated at 25°C for 10 minutes and subsequently spun down at 350xG at 4°C for 5 minutes. The cell pellet was resuspended in 500 μL Accutase (Thermo Fisher, 00–4555–56) and incubated at room temperature for 10 minutes to achieve a single cell suspension^2^. Afterwards, 500 μL DMEM/F12 supplemented with 10% FBS was mixed into the cell suspension, followed by centrifugation at 350xG at 4°C for 5 minutes. After removal of the supernatant, the cell pellet was resuspended in 100 μL 1x PBS and the suspension was passed through a 35 μm cell strainer (Falcon, 352235) to remove cellular aggregates, followed by cell counting using a disposable Neubauer chamber (NanoEnTek, DHC-N01). Cell and gel bead emulsions were generated by loading the cell suspension into the 10X Chromium Controller using the 10x Genomics Single Cell 3’ Library Kit v3.1 (10x Genomics; PN 1000223; PN 1000157; PN 1000213; PN 1000122). The subsequent steps of reverse transcription, cDNA amplification and library preparation were performed according to the manufacturer’s protocol. Note that the incubation at 85°C during reverse transcription was extended to 10 minutes to ensure virus inactivation. The final libraries were pooled (S2 flowcell: up to 8 samples, S4 flowcell: up to 20 samples) and sequenced with the NovaSeq 6000 Sequencing System with either S2 or S4 flow cells (Illumina, paired-end, single-indexing).

^1^Samples that were not immediately prepared for cell encapsulation were resuspended in cryopreservation medium [20% FBS (Gibco, 10500), 10% DMSO (Sigma-Aldrich, D8418), 70% DMEM/F12] and stored at −80°C. Frozen cells were thawed quickly at 37°C, spun down at 350xG at 4°C for 5 minutes, and proceeded with normal processing. In cases where satisfactory cell suspensions were achieved without protease treatment, the samples were filtered through a 20 cell strainer (pluriSelect, 43–50020–03) and directly loaded for encapsulation.

### qRT-PCR

After DTT treatment of the patient samples, 140 μl supernatant was used for the extraction of viral RNA with the QIAmp Viral RNA Mini Kit (QIAGEN, 52904). Quantitative RT-PCR for the E gene of SARS-CoV-2 as published by Corman *et al* (2020) of the extracted RNA in technical triplicates along with three ten-fold dilutions of standardized SARS-CoV-2 genome equivalents was performed using the SuperScript III One-Step RT-PCR Kit (Thermo Fisher, 12574026) and the Roche LightCycler 480.

### Pre-processing and data analysis

The raw 3’ single cell RNA sequencing data were processed using CellRanger version 3.1.0 (10X Genomics). The transcripts were aligned to a customized reference genome in which the SARS-CoV-2 genome (Refseq-ID: NC_045512) was added as an additional chromosome to the human reference genome hg19 (10X genomics, version 3.0.0). An entry summarizing the entire SARS-CoV-2 genome as one “gene” was appended to the hg19 annotation gtf file, and the genome was indexed using “cellranger_mkref.”

After preprocessing, contaminating ambient RNA reads were filtered by using SoupX version 1.2.2 Young & Behjati (2020), https://github.com/constantAmateur/SoupX] and MUC1, MUC5AC, and MUC5B as marker genes. The filtered expression matrices were loaded into Seurat version 3.1.4.9012 (https://github.com/satijalab/seurat), where further filtering was done to remove cells with less than 200 genes expressed or more than 15% mitochondrial transcripts.

An upper cutoff for the number of unique molecular identifiers (UMIs) was manually determined for each sample based on a plot of gene count vs. UMI count with values ranging between 50,000 and 200,000 UMIs. Following QC filtering, the samples were normalized to 10.000 reads, scaled, and centered. For integration, 3,000 shared highly variable genes were identified using Seurat’s “SelectIntegrationFeatures()” function. Integration anchors were identified based on these genes using canonical correlation analysis [CCA, Stuart *et al* (2019)] with 90 dimensions as implemented in the “FindTransferAnchors()” function. The data were then integrated using “IntegrateData()” and scaled again using “ScaleData()”. PCA and UMAP dimension reduction with 90 principal components were performed. A nearest-neighbor graph using the 90 dimensions of the PCA reduction was calculated using “FindNeighbors()”, followed by clustering using “FindClusters()” with a resolution of 0.6. Viral load was calculated on the raw data matrices output by CellRanger. All reads aligning to the SARS-CoV-2 genome per sample were aggregated and divided by the total number of reads in that sample. Multiplication by 10^6^ then yielded the CPM viral load values.

Cell-cell interactions were calculated using CellPhoneDB version 2.1.2 [Efremova *et al*. (2020), https://github.com/Teichlab/cellphonedb] on data subsampled according to the following strategy: in order to decrease the impact of samples with high cell numbers, all samples were randomly down-sampled to the size of the smallest sample. A maximum of 2.000 cells per cell type was kept to reduce calculation times. Transcription factor importance was scored using arboreto 0.1.5 [Moerman *et al* (2019)] and pyscenic 0.10.0 [Aibar *et al* (2017)].

To infer the cluster and lineage relationships between the different epithelial cell types identified, Monocle3 was used [Qiu *et al*. (2017), Cao *et al* (2019)] (https://github.com/cole-trapnell-lab/monocle3). UMAP embeddings and cell clusters generated from Seurat were used as input while trajectory graph learning, and pseudo time measurement through reversed graph embedding were performed with Monocle3. In order to identify genes that are significantly regulated as the cells differentiate along the cell-to-cell distance trajectory, we used differentialGeneTest() function implemented in Monocle2 [Qiu et al, 2017]. This function fits a vector generalized additive model (VGAM) from the VGAM package for each gene in our dataset where the gene expression levels are modeled as a smooth non-linear function of the pseudotime value of each cell. Thereafter, the genes were ranked by the order of their importance based on q-value. For the basal IFN trajectory, we performed a VGAM fit on basal, basal IFN, diff-ciliated & ciliated clusters. The diff-secretory, secretory, FOXN4^+^, diff-ciliated and ciliated cell types were considered when identifying important genes for the FOXN4+ trajectory.

### Statistics

For comparisons between expression values, the Seurat function “FindMarkers()” was used with the “DESeq2” method. Cell type markers were obtained using the “FindAllMarkers()” function with a negative binomial test and the case severity as latent variable. Cell type numbers were compared using Fisher’s exact test. In cases where several samples were aggregated, they were first subsampled to equal cell numbers to avoid dominance of individual samples. Correlation was assessed using Pearson’s R, and significance of correlation was calculated using the corr.test function in R. All tests were two-sided. P-values were corrected using the Benjamini-Hochberg (FDR) method.

### Identification of cell type and state

Epithelial and immune cell types were primarily annotated based on their intermediate expression levels of their respective cell type markers (Extended Data Fig. 1). All previously described major epithelial cells of the conducting airways, including basal, secretory, and ciliated cells, as well as the recently discovered FOXN4^+^ cells and ionocytes were found. In addition, we identified a cell type from the most posterior region of the nasopharynx that we call “squamous epithelial” that is characterized by a strong expression of SPRR genes essential to squamous cell cornification (Figs. 2a and b, Extended Data Fig.1a) [Kalinin et al (2002); Steinert et al (1998)]. Basal, secretory, and ciliated cells were present in different cell states from differentiating to terminally differentiated as shown by their intermediate expression levels of their respective cell type markers (Extended Data Fig. 1), inferred mitotic stages (Extended Data Fig. 2a and 2b), and pseudotime analysis (Extended Data Fig. 3). In particular, diff-secretory and diff-ciliated cells represent the differentiating state, while secretory and ciliated cells are further differentiated. Notably, the FOXN4^+^ cells resemble the transient secretory cell type described as the most SARS-CoV-2 vulnerable bronchial cells in non-infected individuals by virtue of their function as transitionary cell types differentiating from the secretory to ciliated lineage [Lukassen *et al*. (2020)] (Fig. 2a and Extended Data Fig. 3a).

Within the immune cell population, we identified six different cell types, namely macrophages, dendritic cells, mast cells, neutrophils, B and T cells. (Fig 2a and b, Extended Data Fig. 1b, Suppl. Table 1). In the monocyte-derived (MD) fraction, we distinguished resident macrophages (rMa), MD macrophages (moMas), MD dendritic cells (moDCs), and non-resident macrophages (nrMa) [Martinez *et al* (2006), Gren *et al* (2015)], whereas in the T lymphocyte population we identified *CD8^+^* cytotoxic (CTL), lytic *CD8^+^* effector (LyT), T regulatory (Treg), and natural killer T (NKT) cells [Travaglini *et al* (2019)]. The CTL, LyT and NKT cell populations were similar to each other and mainly showed a quantitative difference in the expression of genes rather than a strong difference in characteristic markers. The canonical Treg marker *FOXP3* was not expressed in our dataset. Therefore, Treg specific genes were determined by applying the Seurat FindMarker() function comparing Treg to other T cell gene expression datasets.

Cell type identification was similarly done for samples from nasopharyngeal swabs (NS), bronchial protected specimen brushes (PSB), and bronchioalveolar lavages (BAL) for two critical COVID-19 patients (#32, #62) (Extended Data Fig. 6). We used the same markers as shown in Extended Data Fig. 1 to determine cell types/states. As additional cell types in these samples, we characterized secretory (Secretory *LYPD2*) and ciliated (Ciliated IFN) cell clusters with a heterogeneous expression of ISGs (*IFI6, IFI16, IFI44, IFIT3, IFITM3*). For one particular patient (#62), we identified a “hybrid” cell type that expresses mixed markers for secretory and ciliated cells (*SCGB1A1, MUC5AC, MSMB, TFF3, AGR2, CAPS, SNTN*). Within the immune cell population, we also found natural killer cells (*NCAM1, FCGR3A, KLRD1*) and plasma cells (*CD27, SDC-1, CD79A*).

### Data availability

Due to potential risk of de-identification of pseudonymized RNA sequencing data the raw data will be available under controlled access in the EGA repository upon journal acceptance. In addition, count and metadata tables containing patient ID, sex, age, cell type and QC metrics for each cell will be available at FigShare. All count and metadata will be integrated for further visualization and analysis in Magellan COVID-19 data explorer at https://digital.bihealth.org. For access to data prior to journal acceptance, please contact the corresponding authors.

### Code availability

No custom code was generated or used during the current study.

## ACKNOWLEDGEMENTS

We thank our patients for kindly donating samples and data to this study. The German Bioinformatics Network de.NBI supported this study in technical aspects regarding the efficient processing of sequencing data. We thank Illumina GmbH for financial support via the allocation of reagents and sequencing flow cells as well as Markus Vossmann, Martin Allgaier, Oliver Krätke for the realization of the sequencing runs at the Illumina Solutions Center Berlin. We further wish to thank Intel Germany GmbH for generously donating computational infrastructure. This study was supported by the COVID-19 research program of the Berlin Institute of Health, the German Research Foundation through SFB-TR84 “Innate Immunity of The Lung”, the German Ministry of Education and Research in the framework of the CAPSyS project (Grant 01ZX1304B and 01ZX1604B), the Berlin-University Alliance (BUA), and the European Commission (ESPACE, 874710, Horizon 2020).

## AUTHOR CONTRIBUTIONS

C.C., I.L., L.E.S., and R.E. conceived, designed, and supervised the project.

R.L.C, J.L., F.P., D.W., J.K. and J.L. performed experiments. R.L.C., S.L., S.T., B.P.H., F.P., O.D., L.T., N.I., C.C, I.L., and R.E. analyzed data. D.W., F.K., F.M., H.M-R., A.K., S.S., F.B., C.v.K., M.W., and L.E.S. provided the human specimens and clinical annotation of the patients. A.H., C.G., C.D., and S.L. contributed with discussion of the results. B.T., S.T., S.S, J.E. provided technical and experimental support. S. T., S. S., and J.E. developed Magellan. R.L.C., S.L., S.T., B.P.H., S.L., C.C., L.E.S., I.L., and R.E. wrote the manuscript, all authors read, revised, and approved the manuscript.

## MATERIALS & CORRESPONDENCE

Material requests should be addressed to

Correspondence to

## EXTENDED DATA

**Extended Data Figure 1. Markers used to identify and stratify the different cells of the upper respiratory tract. A**. Dot plot depicting the canonical markers of the epithelial cell types. **B**. Dot plot depicting the markers used to identify the different immune cell types. For details on cell types and markers refer to Methods.

**Extended Data Figure 2. Characterization of *ACE2+* cells from the upper respiratory tract. A**. Proportion of cells in G1, G2/M, or S phase for each cell type. **B**. UMAP depicting cells in either G1, G2/M, or S of a subsample of the entire dataset (10,000 cells, see Methods). **C**. Percentage of *ACE2^+^* cells across control and Influenza B as well as moderate and critical COVID-19 patients. **D**. Composition of *ACE2^+^* cells for all COVID-19 patients in terms of double or triple positive status for *TMPRSS2* and *FURIN*. G1 – Gap I phase, G2 – Gap II phase, M – Mitotic phase, S – Synthesis phase.

**Extended Data Figure 3: Inferred differentiation paths for epithelial cells from the upper respiratory tract. A**. Pseudotime trajectory projected onto a UMAP of selected epithelial cells. Pseudotime values are color-coded. The red dot indicates the start point of the trajectory. Numbers in circles indicate two inferred differentiation paths **B**. Cell density of lung epithelial cell clusters along a cellular trajectory reflecting epithelial cell differentiation. Cells are ordered by pseudotime (cell-to-cell distance metric) as computed by Monocle3. **C**. Expression of specific genes along the cell trajectory. The dots indicate the gene expression of individual cells colored by the epithelial cell type. The green lines approximate expression along the inferred trajectory by polynomial regression fits.

**Extended Data Figure 4: Identified regulons that are activated in ACE2+ cells. A**. Transcription factors ranked according to association strength with *ACE2* expression (for details see Methods). Top 10 hits are highlighted. **B**. Expression profile of the interferon signaling mediators in all cell types. Please note that *IFNG* is strongly expressed in CTL. Expression levels are color-coded, the percentage of cells expressing the respective gene is size-coded.

**Extended Data Figure 5: Interaction of chemokines with their respective receptors comparing controls, to moderate and critical COVID-19 cases**. Expression of chemokines and their respective receptors in the different cellular subpopulations is depicted by dot plots. Connecting lines represent receptor/ligand pairs as obtained from the molecular cell atlas of the human lung [Travaglini *et al*. (2019)] complemented by literature research. Expression levels are grey level coded while the percentage of cells expressing the gene are depicted by the different dot sizes. Red circles show significance of expression when comparing critical and moderate patients as calculated by a two-sided significance test. (n=5 moderate and 6 critical COVID-19 patients, 10 samples each).

**Extended Data Figure 6: UMAP depicting cell types identified from sampling of critical COVID-patients across three different sites of the respiratory system**. Nasopharyngeal swabs (NS), bronchial protected specimen brushes (PSB), and bronchioalveolar lavages (BAL) were taken from two different critical COVID-19 patients (#32 and #62). **A**. All cell types identified are projected on a single UMAP. **B**. Distribution of the different cell types across different sampling sites. **C**. UMAP depicting *ACE2^+^* cells and virus^+^ cells across different sampling sites. Please note that the majority of cell types correspond to the cell types identified in the upper respiratory tract (cf. Fig. 2). For identification and description of the additionally identified cell types including secretory LYPD2, ciliated IFN, basophil, NK and hybrid cells please refer to Methods. **D**. Viral RNA reads within aggregated pseudo-bulks for patient #32 and #62 in counts per million (CPM) for the three sampling sites NS, PSB, and BAL. **E**. Viral genome equivalents/ml of patient #32 cell-free sample as measured by quantitative RT-PCR.

**Extended Data Figure 7: Expression levels of *CCL2, CLL3* and their receptors *CCR1, CCR2*, and *CCR5* in immune cells in the lower respiratory system**. Dot plot depicting the average expression of *CCL3/ CCR1, CCR5* and *CCL2/ CCR2, CCR1* in different immune cells in PSB and BAL samples for patients #32 and #62. Connecting lines represent receptor/ligand pairs. Expression levels are color coded while the percentage of cells expressing the gene are depicted by different dot sizes. PSB - (bronchial) protected specimen brush, BAL - bronchioalveolar lavage.

## SUPPLEMENTARY TABLES

**Suppl. Table 1. Contribution of each cell type from each patient in raw numbers and in percentage for each patient**. Cell type numbers were compared using Fisher’s exact test.

**Suppl. Table 2. Contribution of *ACE2^+^*, *FURIN^+^, TMPRSS2*^+^, cells for cell type from each patient in raw numbers and in percentage for each patient**. Cell type numbers were compared using Fisher’s exact test and p-values as well as odds ratios are provided.

